# Trefoil factor 3 positively associates with IL-25 in patients with Chronic Rhinosinusitis

**DOI:** 10.1101/2020.06.21.20136861

**Authors:** Jorge F. Ortiz-Carpena, Ivy W. Maina, Cailu Lin, Neil N. Patel, Vasiliki Triantafillou, Neil N. Luu, Michael A. Kohanski, Edward C. Kuan, Charles C. L. Tong, John V. Bosso, Nithin D. Adappa, James N. Palmer, Danielle R. Reed, De’Broski R. Herbert, Noam A. Cohen

**Affiliations:** Department of Pathobiology, University of Pennsylvania School of Veterinary Medicine, Philadelphia, PA; Department of Otorhinolaryngology-Head & Neck Surgery, University of Pennsylvania Health System, Philadelphia, Pennsylvania, USA; Monell Chemical Senses Center, Philadelphia, PA; Corporal Michael J. Crescenz VA Medical Center, Philadelphia, PA

## Abstract

**Background:** Trefoil factor family cytokines (TFF1-3) have documented reparative and immunoregulatory effects on mucosal epithelial cells that include: blocking apoptosis, directed cell migration, and regulation of pro-inflammatory cytokine production. TFF1 and TFF3 have been shown to undergo altered expression within sinonasal tissues of patients with various forms of chronic rhinosinusitis including those with or wthout polyp associated disease, but the cellular source(s) of TFF members in CRS remains unclear. To further explore the role of TFF3 in inflammatory sinonasal disease, we sought to determine its expression pattern in immune and non-immune cell types in CRS disease.

**Methods:** Polyp and inferior turbinate tissues were isolated from patients undergoing surgical resection for treatment of CRS. Tissue homogenates and single cell suspensions were subjected to enzyme linked immunosorbent assay (ELISA) and single-cell RNA sequencing. Air liquid interface cultures of sinonasal epithelia were subjected to immunofluorescence (IF) microscopy. Interleukin 25, TFF2 and TFF3 protein levels were compared to sinonasal outcome test scores (SNOT-22) to determine whether levels associated with post-operative outcomes.

**Results:** *TFF3* was broadly expressed in goblet cells, ciliated cells, Tuft cells and T lymphocytes. TFF3 protein levels positively associated with IL-25 in CRSwNP patients. Lastly, TFF3 protein levels positively associated with clinical improvement post-surgery.

**Conclusions:** TFF3 is broadly expressed within multiple epithelial and immune cell lineages in patients with CRSwNP. Protein levels of TFF3 correlate with IL-25 and clinical signs of disease, however TFF3 levels associate with clinical improvement following surgical intervention indicating a potential beneficial role for this reparative cytokine in CRS patients.

## Introduction

Chronic rhinosinusitis (CRS) is a complex upper airway inflammatory disease that affects 16% of the U.S. population^1^. Among CRS patients, some present with a neutrophilic disease dominated by Type 1 cytokines, whereas some individuals develop nasal polyps (CRSwNP) exhibit upper respiratory pathology dominated by Type 2 cytokines (e.g. interleukins 4, 5, 13, 25 and 33) eosinophils, mast cells, basophils and group 2 innate lymphoid cells (ILC2)^2^. While the mechanisms responsible for initiation of CRSwNP disease remain obscure, recent work has shown that in these patients, the major source of the Type 2 polarizing cytokine IL-25 is the Tuft cell/Solitary Chemosensory Cell (SCC)^3^. Tuft cells are a rare epithelial cell lineage that utilize taste receptor genes as sensors to the external microenvironment, where they serve a host protective role by augmenting anti-microbial activity of neighboring cells^4^. Tuft cells are also a major source of immunostimulatory mediators such as cysteinyl leukotrienes and IL-25^5^. Tuft cell derived IL-25 promotes ILC2 expansion and this circuit is enriched in nasal polyp tissues from CRSwNP patients^6^. However, the notion that pathological Type 2 diseases of the airway including allergic asthma and CRSwNP are potentially maladaptive wound healing responses has not been widely considered.

Among known pathways of tissue repair, the Trefoil factor family of reparative cytokines (TFF1, TFF2 and TFF3) are small 6-12kDa cloverleaf-shaped molecules secreted mostly from goblet cells at mucosal barrier surfaces^7^. These cytokines promote resolution of tissue injury through the following: resistance to apoptosis, regulation of cell adhesion, suppression of pro-inflammatory cytokines, and directing cellular migration across denuded basement membrane through a process termed restitution^8^. In the context of CRS, increased gene expression levels for TFF1 and 3 in the sinonasal mucosa has been reported for the Type 1 associated CRSsNP subtype^9^. Moreover, increased TFF3 with concomittant decreased TFF1 gene expression levels in the sinonasal mucosa has been reported for the Type 2 associated CRSwNP subtype^10^. TFFs are expressed in healthy and diseased tissue in human and mouse models and correlate with disease severity in asthma and COPD^11, 12^. In mice, it was shown that TFF2 was necessary for the production of IL-33, which aided the initiation of Type 2 inflammation during allergen exposure and pathogen infection of the lower airways^13^. In addition to its immunoregulatory role(s), TFF3 also has been shown to promote differentiation of beta-tubulin expressing ciliated cells through a mechanism that relied upon epidermal growth factor receptor activation^14^. Despite these insights, there is still a poor understanding of whether TFF members are important for immunoregulation, healing post-surgical intervention, and/or developmental cell lineage sources in CRS patients.

In this study, we used single cell RNA sequencing (sc-RNAseq) to map the cellular expression pattern for TFF genes in CRSwNP patient tissue and determine whether Trefoil protein levels correlated with disease outcome. TFF3 was predominantly expressed in a surprisingly wide array of epithelial and immune cell lineages. Immunostaining of nasal polyp tissues revealed TFF3 was expressed in cells with Tuft cell morphology, which was strikingly consistent with positive correlation between TFF3 and IL-25 levels in mucosal tissues of CRS patients. Combined with the positive association between TFF3 levels and improved clinical outcomes post-surgical intervention for polyp removal, this work indicates that TFF3 may be potentially an important biomarker indicating the extent of pathological Type 2 disease in th context of CRSwNP disease.

## Methods

### Patients and specimen collection

Patients with CRS were recruited from the Department of Otorhinolaryngology – Head and Neck Surgery, Division of Rhinology at the University of Pennsylvania with voluntary informed consent. Patients met the criteria for CRS as defined by the Academy of Otolaryngology—Head and Neck Surgery clinical practice guidelines for adult sinusitis^15^. Sinonasal tissue samples were obtained during routine endoscopic sinus surgery for CRS management after patients had previously failed adequate medical therapy. Patients with a diagnosis of cystic fibrosis, sarcoidosis, immunodeficiency and granulomatosis polyangiitis were excluded. This study was approved by the University of Pennsylvania Institutional Review Board.

### Transcriptome Analysis

Polyp and inferior turbinate (IT) samples from two patients with CRS with nasal polyposis (CRSwNP) were rinsed 3 times with Dulbecco modified Eagle medium containing 100 U/mL penicillin (MilliporeSigma, Burlington, MA), 100 μg/mL streptomycin (MilliporeSigma), and 250 ng/mL amphotericin B (MilliporeSigma). Single cell dissociation was performed by incubating 1-2cm samples in Liberase disruption solution (MilliporeSigma). Digestion was stopped with 5mL of of DMEM+ 10% fetal bovine serum and the digest contents were filtered through a 40-micron cell strainer. The filtrate was centrifuged at 1,200 rpm for 5 minutes and the supernatant was discarded. The cells were resuspended in 5 mL of chilled 0.04% bovine serum albumin (BSA) in PBS and counted with a manual cytometer using trypan blue (Thermo Fisher Scientfic, Waltham, MA). Chilled 0.04% BSA in PBS was added to achieve a target cell concentration of 700-1200 cells/µl. Cell suspensions were loaded on a GemCode Single-Cell Instrument (10x Genomics, Pleasanton, CA) along with reverse transcription reagents, gel beads containing barcoded oligonucleotides and oil to generate single-cell Gel Beads in Emulsion. Using the GemCode Single-Cell 3’ Gel Bead and Library kit (10x Genomics) an scRNA-seq library was prepared. These sequencing libraries were loaded on an Illumina NextSeq500 (Illumina, San Diego, CA) and demultiplexing, barcode processing and single-cell 3’ counting were performed using the Cell Ranger Single-Cell Software Suite (10x Genomics).

### Tissue homogenization

Polyp and IT samples from 16 patients with CRSwNP and 12 patients with CRS without nasal polyposis (CRSsNP) were collected and rinsed as described above. Samples were finely minced in a solution of 1 mL of 1× PBS containing complete Mini Protease Cocktail Inhibitor (Roche, Indianapolis, IN) and homogenized on ice using a Dounce homongenizer. Homogenized samples were transferred to an Eppendorf tube, centrifuged at 14,000 rpm for 5 minutes and the supernatant was transferred to a fresh Eppendorf tube. Samples were stored at −20°C until assayed for TFF3, TFF2, IL-25 and total protein levels, as described below.

### TFF2, TFF3, IL-25 ELISA and total protein quantification

Human TFF2, TFF3 and IL-25 ELISA (R&D Systems, Minneapolis, MN) was performed according to the manufacturer’s instructions with samples of homogenized sinonasal tissue (at 1:5 and 1:15 dilutions). Total protein concentrations for homogenized tissue samples were determined using a bicinchoninic acid protein assay (Thermo Fisher Scientific) in microwell format according to the manufacturer’s instructions and TFF2, TFF3, and IL-25 levels were normalized to total protein levels. Optical density was measured with a microplate reader at wavelengths of 450nm and 570nm for TFF3, TFF2, IL-25 and 562nm for total protein.

### Chart Review

Demographic data, including age and gender, were collected along with disease characteristics (including prior surgeries, pre-and post-operative scores on the validated 22-item sino-nasal outcomes test (SNOT-22^16^ and comorbidities (including prior diagnoses of asthma and allergic rhinitis) (Table 1). Pre-operative SNOT-22 scores were collected 1-4 weeks prior to day of surgery. Post-operative SNOT-22 scores were collected 4-6 weeks after surgery. Delta SNOT-22 scores were calculated as the pre-operative SNOT-22 score minus the post-operative SNOT-22 score. Pre-operative radiographic computerized tomography (CT) scans of the sinuses were also reviewed and scored based on the Lund-Mackay (LM) staging system for CRS^17^.

**Table 1.**
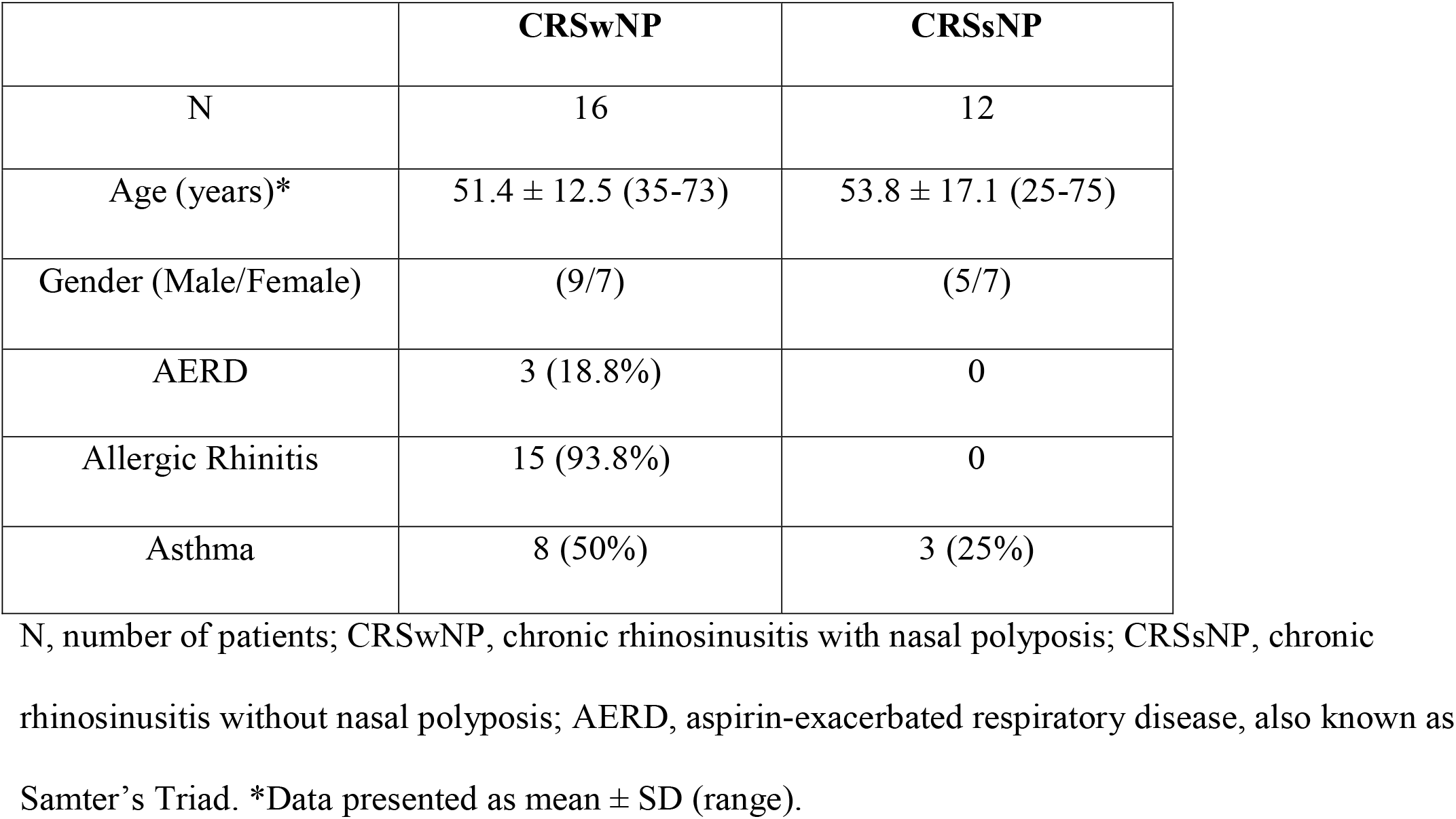
Characteristics of the patients enrolled in the study.

### Analyses

Data were analyzed using Microsoft Excel (Microsoft, Redmond, WA) and Prism (GraphPad Software, La Jolla, CA) software. Linear correlations between TFF3, TFF2, and IL-25 protein levels, and between TFF3 protein levels and SNOT-22 scores (baseline and delta) were assessed using the non-parametric Spearman’s rank correlation test.

Single-cell RNA sequencing results were analyzed using the computational R toolkit Seurat^18^. We processed two scRNA-seq datasets (2 inferior turbinate and polyp samples were merged and filtered, n= 14,013 cells) and created objects with molecular features which are detected in at least 3 cells; each cell had at least 200 molecular features. UMAP plots with single cells were colored by K-nearest neighbor (KNN) clusters and cell types using a ROC test^19^. The top 20 gene markers for each cluster were exported for cell annotation according to the public cell marked database^20^.

## Results

### Among TFFs, TFF3 is selectively enriched in multiple cell populations during CRS disease

Given previous reports that TFF levels are elevated in CRS tissues^9^, we sought to interrogate the lineage identity of TFF expressing cells using single cell RNA-sequencing (scRNA-seq). To this end, we subjected a pool of tissues collected from inferior turbinate and polyp tissue for the generation of single cell suspensions. We prepared sequencing libraries from 15,423 single cells, which were loaded into the Illumina NextSeq 500 Sequencing System (Illumina, San Diego, CA) and processed using the 10x Genomics Software Suite. We explored our scRNA-seq dataset using the computational R toolkit Seurat^18^ to identify cell clusters and differential gene expression within these cell clusters at the transcript level. We identified 13 cell clusters which were annotated by cell type with the top 20 lineage-specific genes per cluster using the CellMarker public database^20^ (Figure 1A). Comparison of cell clusters that showed positive expression for one or more member of the TFF family, we noted that TFF2 showed the most limited expression whereas TFF3 was most broadly expressed (Figure 1B-C). Combined analysis of both inferior turbinate and polyp tissue revealed that TFF1 was a predominant feature of the goblet cell cluster, whereas TFF3 was expressed in clusters identified as goblet cells, ciliated cells, basal cells, and SCC/Ionocytes (Figure 1D). Interestingly, we found overlapping expression between TFF3 and the SCC/tuft cell lineage specific genes *POU2F3* and *TRPM5* (Figure 1C). Consistent with our previous work, the scRNA-seq analysis reveals increased Il25 mRNA expression in polyp tissue^3^ (Figure 1D). Unexpectedly, there was a moderate number of TFF3 expressing cells within the immune system as defined by CD45 (*PTPRC*) and CD11c (*ITGAX*) that are genes expressed in NK/T lymphocytes, monocytes/macrophages, mast cells/basophils and B cells/plasma cells (Figures 1A-C). Combined, these data show that TFF3 is the most predominant member among the TFF family and it is expressed broadly in mature and progenitor epithelial lineages and immune cell populations in patients with CRS disease.

**Figure 1.**
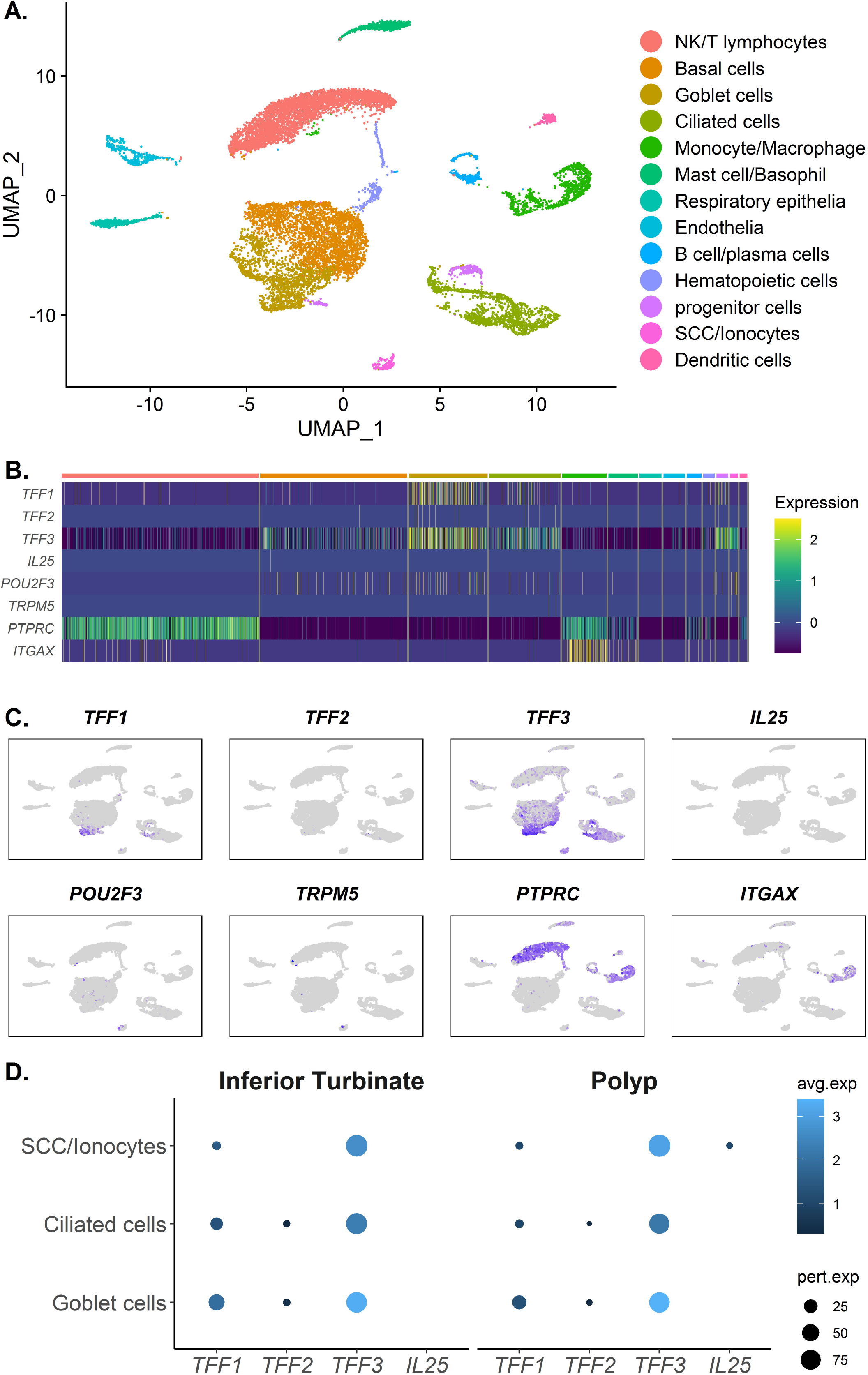
Among Trefoil Proteins, TFF3 is selectively enriched in multiple cells populations during CRS disease. Single-cell RNA sequencing analysis for target genes (e.g., *TFF3*) using the dimension reduction method Uniform Manifold Approximation and Projection (UMAP) from two CRSwNP patient samples. **A**. UMAP plot displaying single cells clustered by KNN clusters and cell types. **B**. Heatmap displaying gene expression across cell clusters defined in (**A**). **C**. Feature plots show expression of *TFF2, TFF3, IL25* and other lineage-defining genes across cell clusters. **D**. Dot plot showing expression of *TFF1, TFF2, TFF3* and *IL25* in Solitary Chemosensory Cells (SCCs), Ciliated Cells, and Goblet Cells from Inferior Turbinate and Polyp tissue. *TFF1*, Trefoil factor 1; *TFF2*, Trefoil factor 2; *TFF3*, Trefoil factor 3; *IL25*, Interleukin 25; *POU2F3*, POU class 2 homeobox 3; *TRPM5*, Transient receptor potential cation channel subfamily M; *PTPRC*, Protein tyrosine phosphatase receptor type C; *ITGAX*, Integrin subunit alpha X.

### TFF3 but not TFF2 correlates with IL-25 production in CRSwNP

Given the central role served by SCC/tuft cell lineages in producing IL-25 to promote Type 2 immunity^21^, we next sought to determine whether TFF3 protein levels in the mucosa of CRS patients correlated with IL-25 levels. Commercially available ELISA kits specific for TFF2, TFF3 and IL-25 were used to compare levels of these cytokines in homogenized tissues from CRSwNP) and non-polyp (CRSsNP) patients. These data were used to generate Spearman correlation coefficients from the linear association of IL-25 protein levels and TFF3 or TFF2 in CRSwNP (Figure 2A) and CRSsNP (Figure 2B). We find that IL-25 protein levels were significantly correlated with TFF3 protein levels in CRSwNP patient samples (Figure 2A: r_s_ = 0.544, p = 0.03). In contrast, there was no significant correlation between IL-25 protein levels and TFF2 protein levels in either CRSwNP (Figure 2A: r_s_ = 0.094, p = 0.72) nor CRSsNP (Figure 2B: r_s_ =0.010, p = 0.97). We also did not find any significant correlation between IL-25 protein levels and TFF3 protein levels in CRSsNP (Figure 2B: r_s_ = 0.055, p = 0.86). Thus, these data suggest that TFF3 but not TFF2 is associated with IL-25 production in CRSwNP.

**Figure 2.**
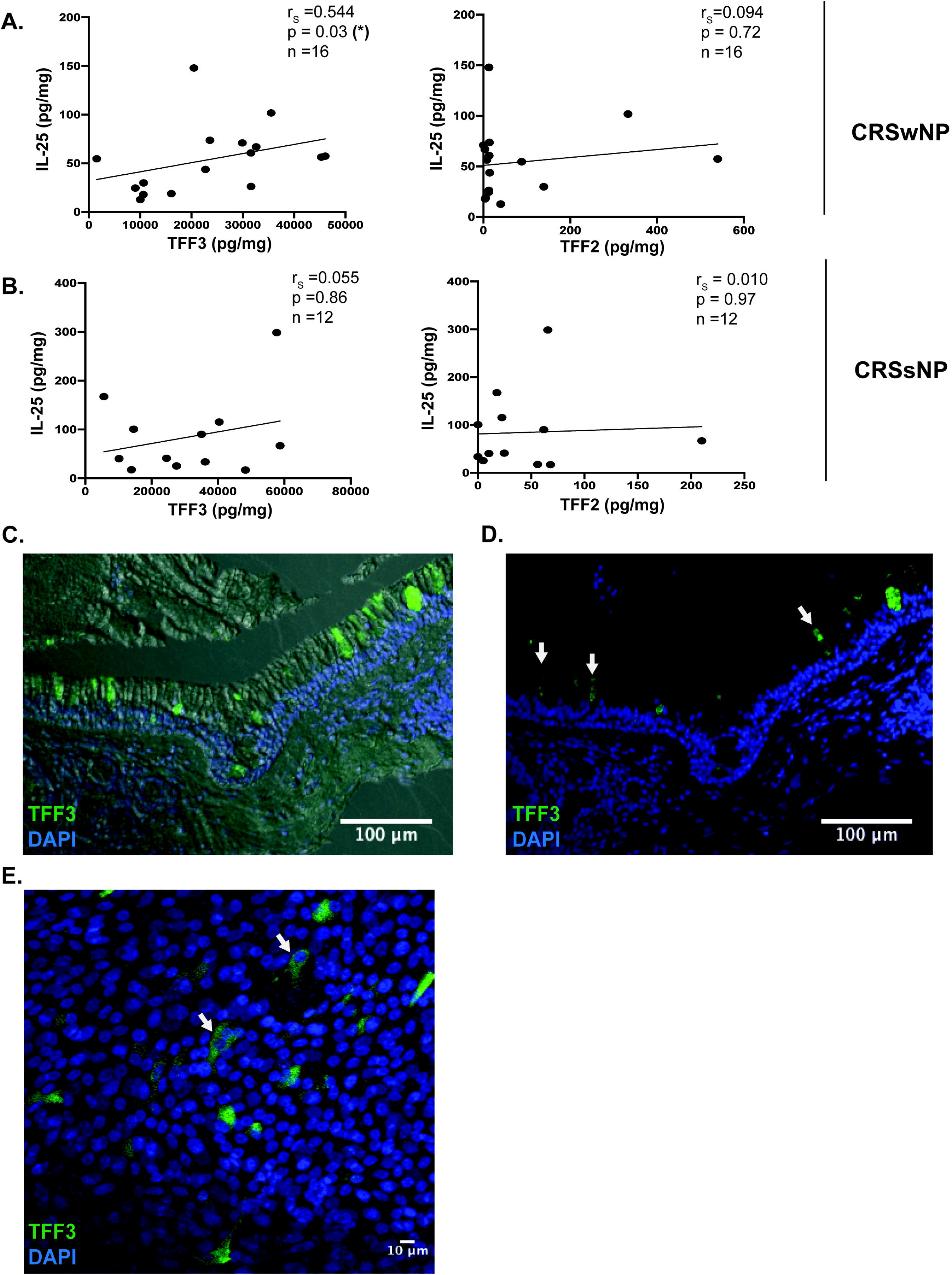
TFF3 but not TFF2 correlates with IL-25 in CRSwNP. **A**. Spearman correlation between protein levels of IL-25 and TFF3 or TFF2 in 16 CRSwNP patient samples measured by ELISA. **B**. Spearman correlation between protein levels of IL-25 and TFF3 or TFF2 in 12 CRSsNP patient samples measured by ELISA. **C**. Differential Interference Contrast (DIC) Imaging of Air-Liquid Interface from sinonasal epithelial cell cultures grown from a CRSwNP patients demonstrating TFF3+ epithelial cells. **D**. Immunofluorescence (IF) Imaging of sinonasal epithelial cell cultures grown from CRSwNP patients demonstrating TFF3 expression in epithelial cells with distinctive SCC/tuft cell-like morphology, indicated by arrowheads. **E**. IF maginification of CRSwNP human nasal epithelium samples.

Next, TFF3-specific immunostaining was performed on air-liquid interface (ALI) cultures of a sinonasal epithelial cell cultures grown from a CRSwNP patients. Evaluation of these tissues using differential interference contrast imaging overlaid upon the fluorescence signal revealed punctate TFF3 staining in cells with apical microvilli and tuft-like cell morphology (Figures 2C-E). Collectively, these data reveal that TFF3 and IL-25 protein levels are closely correlated within the mucosal tissue of CRSwNP patients.

### Impact of TFF3 on clinical markers of disease severity and post-operative tissue repair in CRS disease

Lastly, we asked whether TFF3 correlated with clinical markers of disease severity and post surgical symptom resolution. The Sino-Nasal Outcome Test (SNOT-22) is a patient-reported outcome measure for patients with sinonasal disease with 22 questions designed to assess symptom severity^22^, and is often used to evaluate efficacy of therapeutic intervention^23^. Therefore, we compiled baseline and post-operative (6 month) SNOT-22 scores and calculated the change from baseline to post-operative SNOT-22 scores (deltaSNOT-22) from All CRS patients (n=25), CRSwNP (n=13) and CRSsNP (n=12). We then determined Spearman correlation coefficients from the linear association of TFF3 and baseline SNOT-22 scores (Figures 3A-C) or the change in SNOT-22 scores following surgical intervention (deltaSNOT-22 scores) (Figures 3D-F) in the following groups: 1) all CRS patients (Figures 3A, 3D), 2) CRSwNP (Figures 3B, 3E) and 3) CRSsNP (Figures 3C, 3F). Data show that the association between TFF3 protein levels and baseline SNOT-22 scores (n = 25) was significant for all CRS patients samples (Figure 3A: r_s_ = 0.386, p = 0.05) and CRSwNP patient samples (Figure 3B: r_s_ = 0.558, p = 0.05).

**Figure 3.**
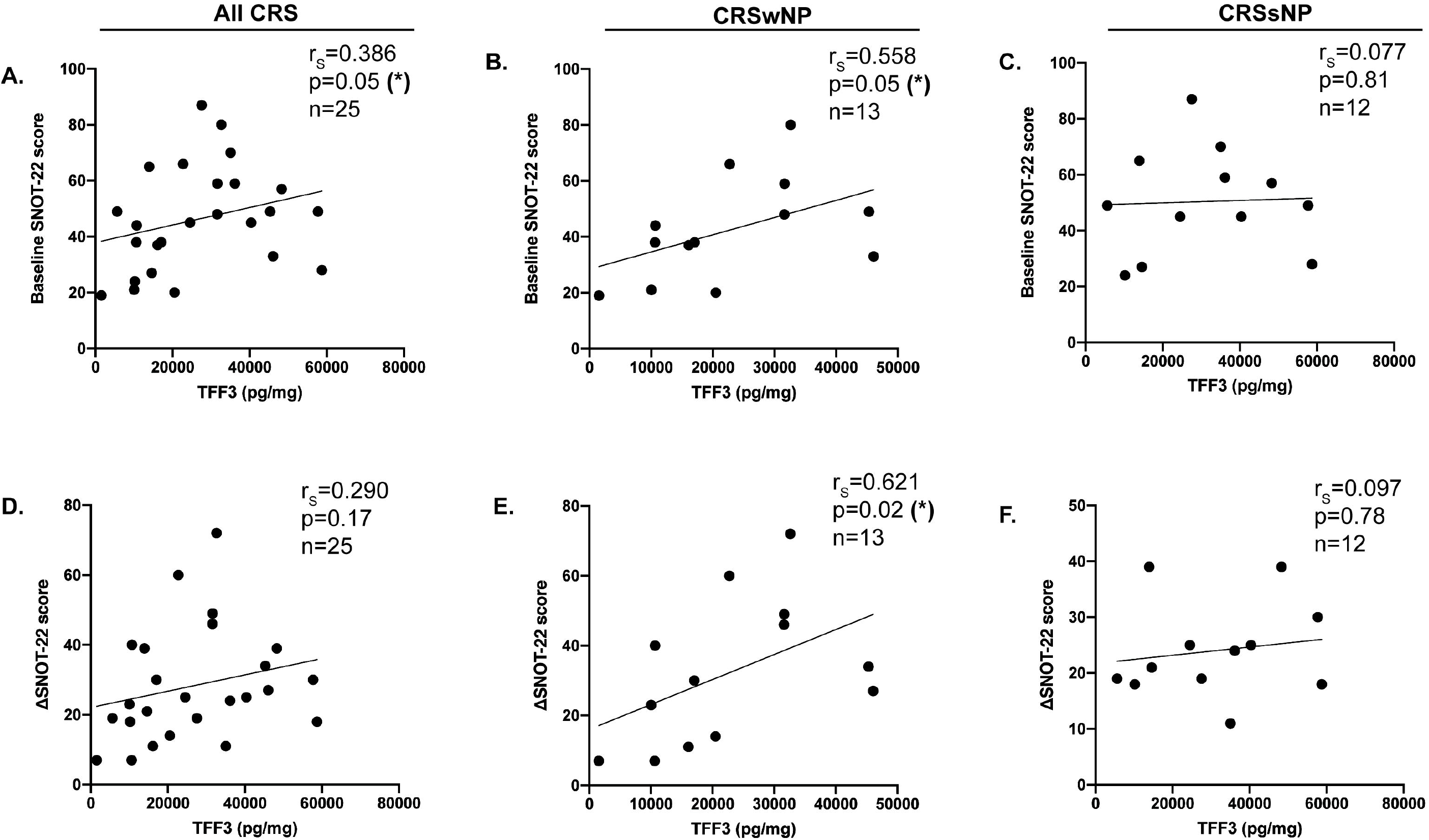
Impact of TFF3 on clinical markers of disease severity and post-operative tissue repair in CRS disease. We performed spearman correlation analyses between Baseline SNOT-22 scores, a clinical marker of disease severity, and TFF3 protein levels in All CRS (**A**), CRSwNP (**B**), and CRSsNP (**C**). Next, we performed spearman correlation analyses between deltaSNOT-22 scores, a metric of the change from pre to post-operative tissue repair, and TFF3 protein levels in All CRS (**D**), CRSwNP (**E**), and CRSsNP (**F**).

Further, when we interrogated the association between TFF3 protein levels and deltaSNOT-22 scores, there was only a significant correlation in CRSwNP patient samples (Figure 3E: r_s_ = 0.621, p = 0.02). There was no significant correlation between TFF3 protein levels and baseline SNOT-22 scores in CRSsNP samples (Figure 3C: r_s_ = 0.077, p = 0.81). Also, there was no significant association between TFF3 and deltaSNOT-22 scores in all CRS patient samples (Figure 3D: r_s_ = 0.290; p = 0.17) or CRSsNP patient samples (Figure 3F: r_s_ = 0.097; p = 0.78). We found no association between TFF3 protein levels and LM score overall or for any sub-group (data not shown). Taken together, these data show that TFF3 correlates with IL-25 production in CRSwNP (Figure 2A) as well as quality of life improvement following sinus surgery (Figure 3E).

## Discussion

CRS is a complex and multi-factorial disease that encompasses a spectrum of immunological phenotypes ranging from Type 1/17 associated neutrophilic inflammation to a Type 2 associated eosinophil dominated response^2^. Moreover, the pathology that accompanies Type 1/17 associated CRS is fibrosis, basement membrane thickening and goblet cell hyperplasia, whereas patients with Type 2 associated CRS develop nasal polyp disease^24^. The underlying causes for CRS remain largely unknown irrespective of the disease subgroup, but prevailing theory posits that chronic defects in mucociliary clearance and/or epithelial repair mechanisms are responsible. This study investigated whether reparative molecules in the Trefoil factor family associate with Type 2 cytokines produced in CRSwNP patients and/or clinical outcomes following surgical intervention. Results from sc-RNAseq analysis show TFF3 was the predominant trefoil factor with a broad expression pattern that included multiple epithelial lineages and immune cells. Moreover, TFF3 protein levels within sinonasal mucosa of CRSwNP patients positively associated with the Type 2 polarizing cytokine IL-25 and greater disease burden as assessed by the SNOT-22 but also correlated with greater improvement following surgery. Taken together, this study warrants further investigation of the underlying mechanistic role(s) served by TFF3 in the pathogenesis of CRS.

Previous work using immunostaining approaches demonstrated that TFF1 and TFF3 are associated with allergic rhinitis and CRSsNP, whereas TFF2 was largely undetectable^9, 25^. In addition, measuring TFF1 and TFF3 mRNA levels in the sinonasal mucosa of CRSwNP patients revealed a significant upregulation of TFF3 gene expression concomittant with downregulation of TFF1 gene expression^10^. TFF1 and TFF3 are known products of goblet cells in both healthy and inflamed sinonasal tissues, TFF3 was also produced by ciliated epithelial cells, and submucosal glandular cells ^25, 26^. While our results are consistent with these conclusions, our sc-RNAseq data additionally show that CRSwNP patients have *TFF3* expression in immune cells (NK/T lymphocytes), basal cells, and SCC/tuft cells. This latter finding is particularly significant because SCC/Tuft cells are major producers of IL-25 in CRSwNP patients and this circuit positively associated with ILC2 expansion^6^. Furthermore, basal cells are the progenitor population that gives rise to SCC/Tuft cells in the lower airway following severe influenza injury^27^. TFF3 immunostaining on air liquid interface cultures of sinonasal epithelium from CRSwNP patients revealed a pyramidal morphology consistent with the distinctive nature of the SCC/Tuft population. Further investigation is certainly warranted to understand the significance of this unexpected TFF3 expression within SCC/Tuft cells and potentially pathogenic lymphocyte subsets.

TFF3 has dynamic effects upon epithelial cells that underlie its diverse roles in wound healing. TFF3 promoted differentiation of ciliated epithelial cells, making it possible that TFF3 may aid in the differentiation of IL-25-producing SCC/Tuft cells from basal cell progenitors in the sinonasal mucosa. Moreover, TFF3 has been shown to inhibit apoptosis and promote cellular proliferation, therefore, it may be possible TFF3 works through autocrine or paracrine mechanisms to sustain SCC/Tuft cell derived IL-25. TFF3 blocks epithelial cell apoptosis through a mechanism that requires interactions with the type 1 membrane receptor LINGO2^28^. While the biological role for LINGO2 is largely unknown, LINGO2 polymorphisms are significantly associated with allergic asthma in humans^29^. Nonetheless, whether a TFF3-LINGO2 interaction works directly or indirectly upon SCC/Tuft cells to promote their expansion, survival and IL-25 production in CRS remains entirely unknown. Given such complexity and the enigmatic nature of CRS in general, a genetically tractable experimental model system will be required to thoroughly investigate this hypothesis.

Having found that TFF3 correlates with IL-25 production in CRSwNP patients, we then asked whether TFF3 also correlated with clinical indicators of disease as defined by SNOT 22 scores. Indeed, there was a positive association between TFF3 and increased SNOT 22. These data could either reflect a inductive role for TFF3 in CRSwNP pathophysiology or that increased TFF3 is an endogenous response intended to resolve areas of damaged tissue, but somehow in this context it is insufficient to do so. Thus, while it remains unclear whether TFF3 is serving a protective or pathogenic role, TFF3 levels in general may be an indicator for inflammation among all CRS endotypes but perhaps more significantly in CRSwNP disease. Overall, our data imply that TFF3 levels in sinonasal tissue biopsies could potentially serve as a biomarker for active CRS and/or good clinical outcomes following nasal polyp resection.

In conclusion, this study demonstrates that the reparative cytokine TFF3 is expressed at the mRNA level within multiple immune and non-immune cell types of patients with CRSwNP disease. Protein levels of TFF3 correlate with IL-25 and clinical signs of disease, which indicates that TFF3 deserves further investigation into its mechanism of action in this debilitating chronic inflammatory disease. A greater understanding of TFF3 in this context also has the potential to reveal important information about the role(s) served by SCC/Tuft cells in human disease.

## Conclusions

TFF3 is emerging as a commonly dysregulated reparative cytokine in human diseases that involve mucosal barrier disruption including allergic asthma, COPD and multiple subtypes of CRS. This study demonstrates that TFF3 expression is broadly upregulated in inflamed sinonasal tissue and correlates with IL-25 and clinical markers of disease suggesting that nasal polyp formation is associated with dysregulated epithelial repair pathways. Understanding the mechanisms in which TFF3 signals in the epithelium through receptor mediated pathways is critical for pursuit of TFF3 proteins as clinically relevant therapeutic targets in humans given the promise shown in animal models^30, 31^.

## Data Availability

The data that support the findings of this study are available from the corresponding author, NAC, upon reasonable request.

## Acknowledgements

Funding sources for the study: National Institutes of Health (NIH), R01DC013588-04S2 to I. W. M., GM083204□08A1, UO1AI125940, and R01AI095289 to D. R. H.; R01DC013588 to N. A. C. The content is solely the responsibility of the authors and does not necessarily represent the offices views of the NIH.

## References

1. Blackwell DL, Lucas JW, and Clarke TC. Summary health statistics for U.S. adults: national health interview survey, 2012. Vital Health Stat 10 2014:1–161.

2. Cao PP, Wang ZC, Schleimer RP, et al. Pathophysiologic mechanisms of chronic rhinosinusitis and their roles in emerging disease endotypes. Ann Allergy Asthma Immunol 2019; 122:33–40.

3. Kohanski MA, Workman AD, Patel NN, et al. Solitary chemosensory cells are a primary epithelial source of IL-25 in patients with chronic rhinosinusitis with nasal polyps. J Allergy Clin Immunol 2018; 142:460–469 e467.

4. Lee RJ, Kofonow JM, Rosen PL, et al. Bitter and sweet taste receptors regulate human upper respiratory innate immunity. J Clin Invest 2014; 124:1393–1405.

5. Bankova LG, Dwyer DF, Yoshimoto E, et al. The cysteinyl leukotriene 3 receptor regulates expansion of IL-25-producing airway brush cells leading to type 2 inflammation. Sci Immunol 2018; 3.

6. Patel NN, Kohanski MA, Maina IW, et al. Solitary chemosensory cells producing interleukin-25 and group-2 innate lymphoid cells are enriched in chronic rhinosinusitis with nasal polyps. Int Forum Allergy Rhinol 2018.

7. Taupin D, and Podolsky DK. Trefoil factors: initiators of mucosal healing. Nat Rev Mol Cell Biol 2003; 4:721–732.

8. Kjellev S. The trefoil factor family - small peptides with multiple functionalities. Cell Mol Life Sci 2009; 66:1350–1369.

9. Li P, and Turner JH. Chronic rhinosinusitis without nasal polyps is associated with increased expression of trefoil factor family peptides. Int Forum Allergy Rhinol 2014; 4:571–576.

10. Mihalj M, Bujak M, Butkovic J, et al. Differential Expression of TFF1 and TFF3 in Patients Suffering from Chronic Rhinosinusitis with Nasal Polyposis. Int J Mol Sci 2019; 20.

11. Viby NE, Pedersen L, Lund TK, et al. Trefoil factor peptides in serum and sputum from subjects with asthma and COPD. Clin Respir J 2015; 9:322–329.

12. Viby NE, Nexo E, Kissow H, et al. Trefoil factors (TFFs) are increased in bronchioalveolar lavage fluid from patients with chronic obstructive lung disease (COPD). Peptides 2015; 63:90–95.

13. Wills-Karp M, Rani R, Dienger K, et al. Trefoil factor 2 rapidly induces interleukin 33 to promote type 2 immunity during allergic asthma and hookworm infection. J Exp Med 2012; 209:607–622.

14. LeSimple P, van Seuningen I, Buisine MP, et al. Trefoil factor family 3 peptide promotes human airway epithelial ciliated cell differentiation. Am J Respir Cell Mol Biol 2007; 36:296–303.

15. Rosenfeld RM, Piccirillo JF, Chandrasekhar SS, et al. Clinical practice guideline (update): adult sinusitis. Otolaryngol Head Neck Surg 2015; 152:S1–S39.

16. Hopkins C, Gillett S, Slack R, et al. Psychometric validity of the 22-item Sinonasal Outcome Test. Clin Otolaryngol 2009; 34:447–454.

17. Hopkins C, Browne JP, Slack R, et al. The Lund-Mackay staging system for chronic rhinosinusitis: how is it used and what does it predict? Otolaryngol Head Neck Surg 2007; 137:555–561.

18. Butler A, Hoffman P, Smibert P, et al. Integrating single-cell transcriptomic data across different conditions, technologies, and species. Nat Biotechnol 2018; 36:411–420.

19. Parodi S, Muselli M, Fontana V, et al. ROC curves are a suitable and flexible tool for the analysis of gene expression profiles. Cytogenet Genome Res 2003; 101:90–91.

20. Zhang X, Lan Y, Xu J, et al. CellMarker: a manually curated resource of cell markers in human and mouse. Nucleic Acids Res 2019; 47:D721–D728.

21. Schneider C, O’Leary CE, and Locksley RM. Regulation of immune responses by tuft cells. Nat Rev Immunol 2019; 19:584–593.

22. Piccirillo JF, Merritt MG, Jr., and Richards ML. Psychometric and clinimetric validity of the 20-Item Sino-Nasal Outcome Test (SNOT-20). Otolaryngol Head Neck Surg 2002; 126:41–47.

23. Kennedy JL, Hubbard MA, Huyett P, et al. Sino-nasal outcome test (SNOT-22): a predictor of postsurgical improvement in patients with chronic sinusitis. Ann Allergy Asthma Immunol 2013; 111:246–251 e242.

24. Schleimer RP. Immunopathogenesis of Chronic Rhinosinusitis and Nasal Polyposis. Annu Rev Pathol 2017; 12:331–357.

25. Miyahara N, Ishino T, Kono T, et al. Expression of Trefoil factor family peptides in the nasal allergic mucosa. Rhinology 2012; 50:408–416.

26. dos Santos Silva E, Ulrich M, Doring G, et al. Trefoil factor family domain peptides in the human respiratory tract. J Pathol 2000; 190:133–142.

27. Rane CK, Jackson SR, Pastore CF, et al. Development of solitary chemosensory cells in the distal lung after severe influenza injury. Am J Physiol Lung Cell Mol Physiol 2019; 316:L1141–L1149.

28. Belle NM, Ji Y, Herbine K, et al. TFF3 interacts with LINGO2 to regulate EGFR activation for protection against colitis and gastrointestinal helminths. Nat Commun 2019; 10:4408.

29. Berube JC, Gaudreault N, Lavoie-Charland E, et al. Identification of Susceptibility Genes of Adult Asthma in French Canadian Women. Can Respir J 2016; 2016:3564341.

30. Cook GA, Thim L, Yeomans ND, et al. Oral human spasmolytic polypeptide protects against aspirin-induced gastric injury in rats. J Gastroenterol Hepatol 1998; 13:363–370.

31. Babyatsky MW, deBeaumont M, Thim L, et al. Oral trefoil peptides protect against ethanol- and indomethacin-induced gastric injury in rats. Gastroenterology 1996; 110:489–497.

